# Aerosol SARS-CoV-2 in hospitals and long-term care homes during the COVID-19 pandemic

**DOI:** 10.1101/2021.05.31.21257841

**Authors:** Gary Mallach, Samantha B. Kasloff, Tom Kovesi, Anand Kumar, Ryan Kulka, Jay Krishnan, Benoit Robert, Michaeline McGuinty, Sophia den Otter-Moore, Bashour Yazji, Todd Cutts

## Abstract

**Background:** Few studies have quantified aerosol concentrations of SARS-CoV-2 in hospitals and long-term care homes, and fewer still have examined samples for viability. This information is needed to clarify transmission risks beyond close contact.

**Methods:** We deployed particulate air samplers in rooms with COVID-19 positive patients in hospital ward and ICU rooms, rooms in long-term care homes experiencing outbreaks, and a correctional facility experiencing an outbreak. Samplers were placed between 2 and 3 meters from the patient. Aerosol (small liquid particles suspended in air) samples were collected onto gelatin filters by Ultrasonic Personal Air Samplers (UPAS) fitted with <2.5µm (micrometer) and <10 µm size-selective inlets operated for 16 hours (total 1.92m^3^), and with a Coriolis Biosampler over 10 minutes (total 1.5m^3^). Samples were assayed for viable SARS-CoV-2 virus and for the viral genome by multiplex PCR using the E and N protein target sequences. We validated the sampling methods by inoculating gelatin filters with viable vesicular stomatitis virus (VSV), and with three concentrations of viable SARS-CoV-2, operating personal samplers for 16hrs, and quantifying viable virus recovery by TCID_50_ assay.

**Results:** In total, 138 samples were collected from 99 rooms. RNA samples were positive in 9.1% (6/66) of samples obtained with the UPAS 2.5µm samplers, 13.5% (7/52) with the UPAS 10µm samplers, and 10.0% (2/20) samples obtained with the Coriolis samplers. Culturable virus was not recovered in any samples. Viral RNA was detected in 10.9% of the rooms sampled. There was no significant difference in viral RNA recovery between the different room locations or samplers. Method development experiments indicated minimal loss of SARS-CoV-2 viability via the personal air sampler operation.

**Key Findings:** Although a subset of aerosol samples exhibited detectable SARS-CoV-2 RNA at low titres, the presence of viable SARS-CoV-2 virus in aerosols appears to be infrequent at >2m distance.

## Background

SARS-CoV-2, the virus that causes COVID-19, predominantly spreads during close contact between shedding individuals and susceptible persons. The virus spreads mainly by inhalation of small respiratory particles, including from asymptomatic or pre-symptomatic individuals.[1-3] Transmission is most likely to occur during close, sustained contact in indoor environments with a person who is in the early stages of infection, where viral concentrations in the air are highest.[4-6] Once expelled, larger respiratory droplets stay aloft for relatively short periods of time, and typically travel short distances (0 to 2m) while smaller particles (aerosols) stay aloft for longer and are able to cover larger distances.[7] Although transmission during close contact is believed to predominate, aerosol transmission has been reported to occur beyond close contact (e.g., greater than 2m), when infectious respiratory particles are able to accumulate to levels that can cause infection, such as in indoor environments with inadequate ventilation or filtration.[1,8] Actions such as coughing, talking and breathing generate respiratory particles of various sizes, including high numbers of particles smaller than 5-10µm in diameter.[5,9,10] Particles up to 100µm may remain suspended in aerosols, accumulating and spreading through an indoor space, again particularly in the absence of adequate ventilation or filtration.[7] Particles less than 5-10µm in diameter, which may remain airborne for extended periods,[11-13] have been shown to contain infectious particles of SARS, MERS, and H1N1.[14] Indeed, tiny aerosolized particles <1µm may remain airborne for upwards of 12 hours.[15] In addition to aerosols, fomites may contribute to transmission, as the virus can remain viable on various surfaces for prolonged periods.[16-18] Additional data specific to SARS-CoV-2 will help address uncertainties about viral concentrations in aerosols, and whether aerosolized virus is infectious.[13,15,19,20]

Evidence from outbreak investigations supports the view that aerosol transmission beyond close contact occurs in certain circumstances (e.g., when ventilation is inadequate); the frequency of this transmission remains unclear, though it is likely to play a role in super-spreading events.[8,21,22] Moreover, modelling studies show that even during close contact interactions, aerosols, rather than larger ballistic droplets, drive viral exposures, as droplets are relatively unlikely to land on a mucus membrane, compared to the higher likelihood of inhaling aerosols.[23]

Research studies mainly rely on molecular detection of the SARS-CoV-2 genome to determine viral presence in aerosol samples. However, genome (RNA) presence alone is often seen as insufficient evidence that aerosol transmission risk is present, as it does not indicate the presence of viable virus. [24-26] In patient swabs, viable SARS-CoV-2 virus is generally detected only within eight days of symptom onset, while viral RNA persists long beyond infectivity.[24,27] As such, to determine viral infectivity, culturing samples in susceptible cells or animal hosts is needed. Persons that are infected with variants such as B.1.1.7 may shed virus for longer, contributing to higher rates of transmission.[28,29]

Although cellular assays are required to establish viral infectivity, air sampling campaigns using size-selective inlets have rarely employed these methods for SARS-CoV-2, given technical challenges in viral sampling.[13] Several hospital air monitoring studies found the genome of SARS-CoV-2, without culturing the virus. [11,20,30] Sampling using a size selective inlet and outfitted with a gelatin filter, Liu et al. found that the highest concentration of viral RNA was present in the size fraction <2.5µm, which could remain suspended over long periods.[11] Likewise, SARS-CoV-2 RNA has been found on surfaces far from infected individuals, including on exhaust outlets and ceiling fans where only fine, aerosolized particles would be likely to reach.[31] These studies indicate that RNA is present in airborne particles.[13,25] To determine infectiousness, Lednicky et. al. measured virus concentrations inside the car of a person with COVID-19. They sampled in multiple size fractions and cultured for viability, finding viable virus present only in the 0.25-0.5μm size fraction, which also had the highest concentration of viral RNA.[12] It is possible that larger size fractions were preferentially filtered out by the vehicle’s ventilation system.

Lab-based studies generated viable aerosols of <5µm of SARS-CoV-2 that had a half-life of greater than one hour while airborne.[18] Once airborne, virus viability is reduced by several environmental pressures, including sunlight, temperature, and relative humidity, while ventilation and filtration reduce risks of transmission beyond close contact.[32]

The size of a virus-containing particle may also affect infectivity. One study demonstrated that macaques infected by the aerosol route had more severe disease compared to the intratracheal/intranasal route.[33] Submicron particles are able to penetrate more deeply into the lungs, depositing in the alveolar region where immune responses may be evaded.[15] Also, in the alveolar region the ACE-2 receptors that the SARS-CoV-2 virus binds to are more accessible, possibly increasing the likelihood of infection.[15,19] This underlines the importance of sampling for viable virus in fine size fractions.

Organizations including the WHO, REHVA (the Federation of European Heating, Ventilation and Air Conditioning Associations) and ASHRAE (the American Society of Heating, Ventilating, and Air-Conditioning Engineers) have recognized the potential hazard of aerosol transmission indoors.[4,5,34] As such, expert organizations are recommending COVID-19 specific control measures, including increased ventilation rates, avoiding air recirculation, using air cleaning and disinfection devices, and reducing the number of occupants.[4,34,35]

Our study objective was to quantify the concentration of SARS-CoV-2 RNA and viable virus in aerosols collected greater than two meters from patients with COVID-19 infection, specifically in the <2.5um and <10um size fractions. In order to examine the presence of SARS-CoV-2 in a wide range of high-risk environments, we conducted air monitoring in hospital rooms, long-term care facility rooms, penitentiary cells and personal residences housing people with recently diagnosed, active COVID-19 disease.

## Methods and Materials

### Aerosol field sampling

Sampling was conducted adjacent to COVID-19 confirmed-positive patients in hospital rooms (ICU and medical ward beds), residential homes, exhaust (return) air ducts drawing air from inside penitentiary cells and living areas, and in long term care home resident rooms. Respiratory particle samples were collected using Ultrasonic Personal Aerosol Samplers (UPAS, Access Sensor Technologies, Fort Collins, USA) operated at 2LPM (Liters per minute)(filter face velocity of 0.031 m/s), with either a 2.5µm or 10µm size selective inlet to exclude larger particles or droplets. The UPAS filter cartridge was loaded with a sterile 37mm gelatin filter (12602-37-ALK, Sartorius, Göttingen, DE) to preserve the integrity and viability of virus containing particles. UPAS samplers were placed 2 to 3 m from the patient’s head for a 16-hour period sampling a total of 1.92m^3^ of air. Prior laboratory testing indicated that longer sampling duration (larger air sample volumes) could lead to cracking of the gelatin filter membrane. Filters were pre-loaded into the samplers in a HEPA filtered biological safety cabinet and sealed in a Ziploc bag prior to sampling. Field and laboratory blanks were deployed to ensure that samples were not contaminated. Within 2 hours post-collection, gelatin filters were removed from sampler inlets with sterile forceps, dissolved into 2mL of pre-warmed (37°C) Viral Transport Medium (VTM) (HBSS, FBS, Gentamycin and Amphoteracin B) to maintain viability, and kept at 4°C until analyzed.[36,37]

In a subset of locations, we also deployed a Coriolis µ Biosampler (Montigny-le-Bretonneux, France) at an inlet velocity of 150 LPM for 10 minutes (1.5m^3^ sample volume). Coriolis air samples were collected into sterile cones containing 5mL of VTM, which was reduced to 3mL by evaporation during sample collection. The Coriolis sampler collects particles larger than 0.5μm in size, with no specified cutoff. Clinician partners working in the respective institutions deployed instruments, pre-programmed to run at a convenient time, when patients would remain in their rooms.

To ensure that no contaminated instrumentation was re-deployed, after sampling the UPAS and filter cartridges were thoroughly cleaned and sterilized with 70% ethanol and wiped before being used again. Prior to loading the gelatin filters, the UPAS filter cartridges and size selective inlets were washed in D.I. water, dipped in 70% ethanol, and air-dried before assembly. All samples were collected between September 22, 2020 and January 25, 2021.

### Aerosol sample processing

Samples taken within the Winnipeg region were taken to the NML (National Microbiology Laboratory, Winnipeg) within 2 hours post sampling, while samples from the Ottawa area were shipped overnight to the NML in 2mLs of VTM at 4°C. VTM was pre-warmed at 37°C and incubated for 10 minutes to dissolve the gelatin filters. Of the resulting solution, 500ul was aspirated into 6 well tissue culture plates for safety testing to detect viable virus, and 140ul supernatant was used for viral genome detection using the QIAmp Viral RNA minikit, as per the manufactures protocol. In control trials, we found that keeping the dissolved filter at 4°C overnight did not reduce viability or nucleic acid detection, compared with immediate processing.

### Cell culture

African green monkey VeroE6 cells (ATCC CRL 1586; American Type Culture Collection, Manassas, VA, United States) were maintained at 37°C+5% CO_2_ in Cell Culture Medium (CCM) consisting of Dulbecco’s modified Eagle cell culture medium (DMEM; Hyclone SH3024302) supplemented with 10% Fetal Bovine serum (FBS; Gibco 12484028) and 10 units per ml of Penicillin/Streptomycin (PS, Gibco 10378016). Medium for virus cultures (VCM) consisted of DMEM supplemented with 2% FBS and 10 units per ml of PS.

### Virus viability and titration

VeroE6 cells were seeded the day prior in 96-well plates to attain 80% confluence on the day of titrations.[38] Media from the previously seeded plates was aspirated and replaced with 150ul of fresh VCM prior to the addition of the sample inoculum. VTM containing dissolved sample filters was added to dilution blocks and 10-fold serially diluted in VCM where 50 ul was added to plates in replicates of 5 per dilution series. Plates were incubated at 37°C +5% CO_2_ for 5-7 days and examined for CPE where virus titers were calculated.[38]

For qualitative detection of viable virus present (safety testing), media from previously seeded 6-well plates was aspirated and 4 mls of fresh VCM was added. To this, 500ul of sample was added and incubated at 37°C +5% CO_2_ for 5-7 days. Wells were examined, compared to a negative control for CPE, and scored positive or negative based on evident CPE in the cell monolayer.

### Method validation using surrogate virus (VSV)

We evaluated the ability of the gelatin filters to maintain virus viability following a 16-hour sampling period. Vesicular stomatitis virus (VSV) was diluted to 10^5^-10^6^TCID_50_ /ml in a tripartite soil load and spotted onto gelatin filters over 5 different spots in 2ul volumes.[39] Three filters were used as a positive control and processed immediately, three others were kept overnight in the Biosafety Cabinet (BSC), and three gelatin filters were placed in a UPAS sampler where it was left running for 16 hours. The following morning, we inoculated an additional 3 gelatin filters and set a sampling time for 4 hours. After sampling times were complete, 2mLs of pre-warmed VCM was added to gelatin filters and incubated for 10 minutes, centrifuged and tittered.

### Method validation using SARS-C0V-2

To determine technical limits of detection of viable SARS-CoV-2 using UPAS sampling, and to correlate viable particles with genomic detection, we spiked gelatin filters with 10ul of SARS-CoV-2 diluted to 10^3^, 10^4^, or 10^5^ TCID_50_ units/mL over five different spots as described above (3 filters per each of the three dilutions). After a brief drying period under a biosafety cabinet, filters were loaded into the UPAS and run under typical conditions (16hrs at 2 LPM). The third corresponding filter for each solution was processed at time of UPAS run start, to be compared with the sampled filters to determine the degree that viability decreases during sampling. Filters were then dissolved into 2 mL of pre-warmed DMEM, and qualitative isolation and quantitative end-point titration was determined in Vero E6 cells. Concentrations were selected to reflect the range of viable aerosol SARS-CoV-2 concentrations previously reported in a hospital room in Florida.[40]

### Molecular viral load (qRT-PCR)

Molecular viral load was determined by qRT-PCR (QuantStudio 5, Applied Biosytems, USA) using the Taq Path One-Step multiplex mix (Applied Biosystems A28522) with primers and probes targeting the SARS-CoV-2 nucleocapsid (N)[41] gene and envelope (E) protein (table 1).[42] Thermal cycling conditions were 53°C for 10 min for reverse transcription, followed by 95°C for 2 min and then 40 cycles of 95 °C for 2s, 60°C for 30s. Values are reported as log genome equivalents per mL based on cycle threshold (Ct) values obtained with a standard curve of known concentration of viral RNA genome containing both the E and N region.

**Table 1:**
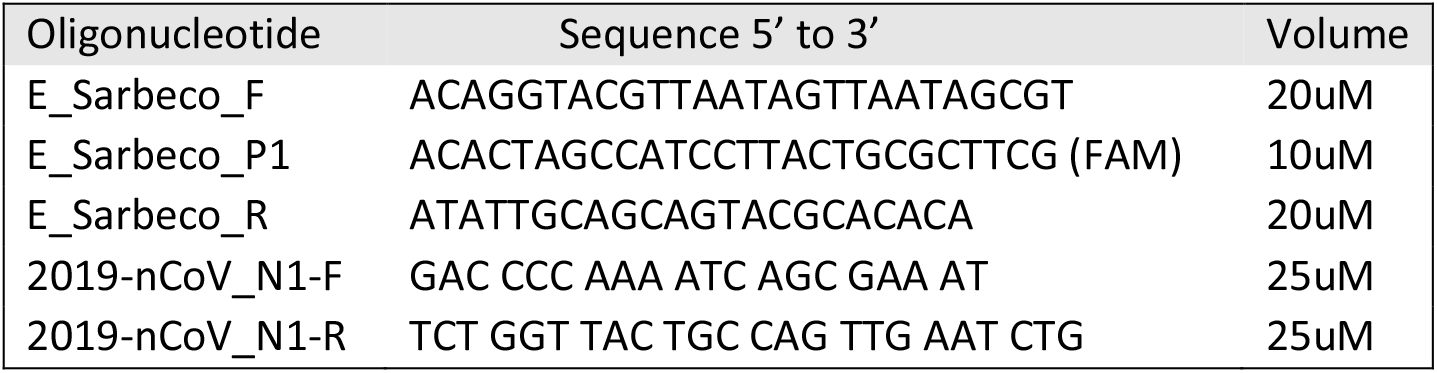
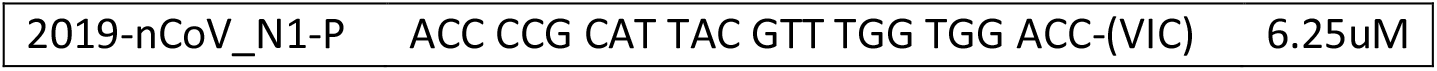
SARS-CoV-2 qRT-PCR primers and probes.

We considered the E protein concentrations as more reliable than N, because the primer probe set is much more sensitive than the N. Samples with a Ct values below 36 for either gene were deemed to be RNA positive. If Ct values were above 36, samples were deemed positive only if detection was confirmed on both the E and N protein target PCRs, with Ct values up to 40.

### Statistical analysis

Data was tabulated in Excel spreadsheets and analyzed using SPSS version 27 (IBM corporation). One sample had a borderline Ct, and was included as a “positive” sample in the analyses. Descriptive statistics were performed. Differences in proportions between groups were evaluated using Chi-square for large tables. Predictors of viral RNA detection were analyzed using Binomial Logistic Regression. As it was unclear which sized particles were most likely to contain viral or viral RNA, if any sampler in a given room was positive for virus, the room was considered to have aerosol virus/viral RNA. Differences between room types for continuous variables were evaluated using One-Way Analysis of Variance. Probability values (p) less than 0.05 were considered statistically significant.

### Ethics

The following Research Ethics Boards were engaged prior to sampling: Health Canada/Public Health Agency of Canada, Children’s Hospital of Eastern Ontario; The Ottawa Hospital; and the Winnipeg Hospital. The boards each determined that formal approval was not required for this study, because sampling was environmental in nature, we did not propose to collect personal information such as time of symptom onset or current symptomatology, and given the urgent need for this data. Moreover, as personal information was not being collected, the boards felt that patient consent was not required.

## Results

### Method Validation Experiments

#### VSV method validation

Results of the VSV experiment are shown in figure 1. Of 5 logs of viable VSV inoculated onto the surface of the gelatin filters, 3 logs of viable virus remained after both 4 hours and 16 hours of drying without any air movement. For the filters that underwent 16 hours of air sampling, 3.5 logs of viable VSV remained, demonstrating that drawing air through the filter did not further reduce viability. It was thus determined that 16 hours would be the optimal sample duration, reflecting the maximum volume of air that could be drawn through the gelatin filter without leading to excessive drying and cracking, or other damage to the gelatin filter.

**Figure 1:**
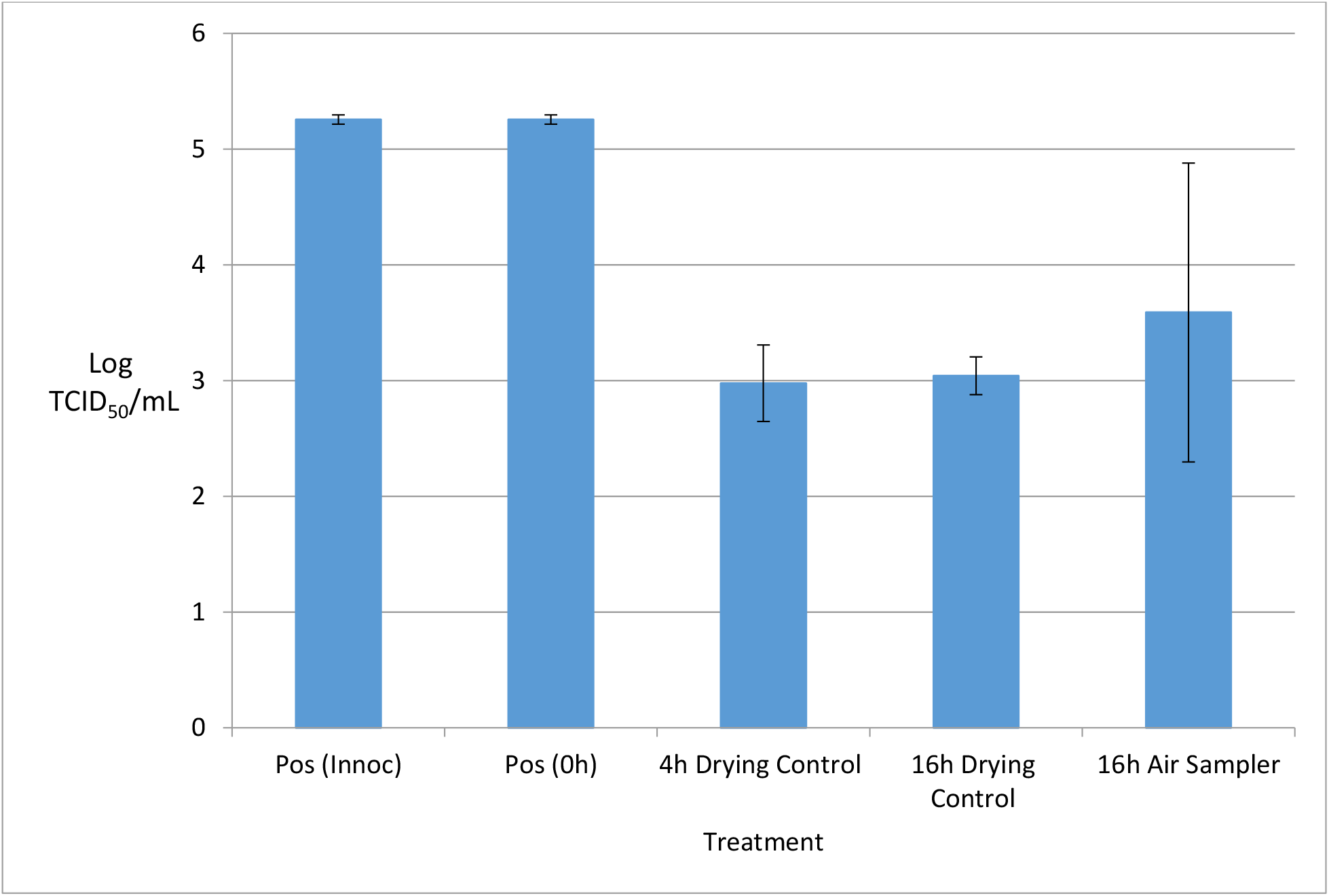
Impact of air sampling on VSV recovery from gelatin filters.

### SARS-CoV-2 method validation

Figure 2 shows that the amount of viable SARS-CoV-2 virus recovered from the gelatin filters was only minimally impacted by sampling in the UPAS for 16 hours (∼2m^3^ sample volume). Similarly, there was minimal loss of viable SARS-CoV-2 when comparing filters processed immediately after inoculation versus those held in VTM at 4°C overnight (Figure 2). Therefore, we are confident in the ability of the UPAS to maintain viability of SARS-CoV-2 once collected into the sampler even at low titers of virus. Furthermore, we compared recovery from samples either processed immediately after dissolving or after dissolved media was placed at 4°C overnight and found no substantial difference.

**Figure 2:**
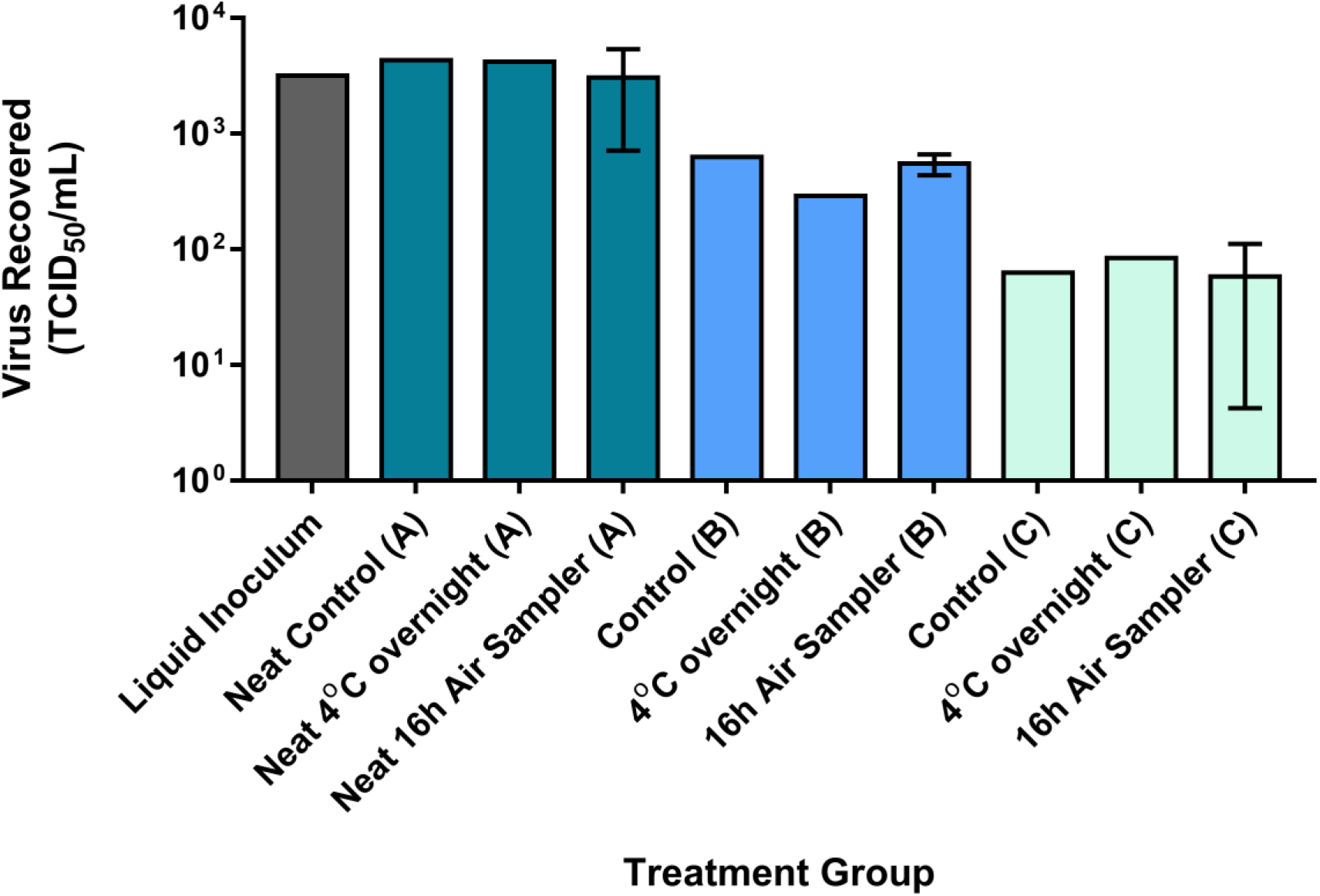
Impact of air sampling on SARS-CoV-2 recovery from gelatin filters. Inocula (A), (B) and (C) were intended to consist of approximately 3.5, 2.5, and 1.5 Log TCID50 of SARS-CoV-2/mL, respectively, with (A) undiluted (neat), and (B) and (C) diluted from the initial Liquid Inoculum. Following one hour of drying, inoculated filters were either placed in UPAS units for 16 hours (Air Sampler), dissolved in VTM and processed immediately (Control), or dissolved at placed at 4°C overnight prior to processing (4°C). Results represent viable virus recovered in VeroE6 cells by TCID_50_ assay. Whiskers represent standard deviation of replicate samples.

Interestingly, SARS-CoV-2 maintained much more viability in the experiments compared to VSV, with the latter undergoing a reduction of over 90% over the sampling period. As such, method development experiments are required to quantify the ability of this sampling method to maintain viability for other viruses of concern.

### Air sampling for SARS-CoV-2

Samples were collected from 99 rooms located in ICUs, hospital ward rooms, rooms in long-term care facilities and at a correctional institute, and a total of 138 samples were obtained. Of the samples, 23 (16.7%) were obtained in ICU rooms, 92 (66.7%) from hospital ward rooms, 15 (10.9%) from rooms in long-term care facilities, and 8 (5.8%) in the correctional institute. 66 samples (47.8%) were obtained using the UPAS 2.5µm sampler, 52 (37.7%) using the UPAS 10µm sampler, and 20 (14.5%) with the Coriolis sampler. In accordance with our procedure, data on patient demographics was not obtained, beyond the fact that all had SARS-CoV-2 infection confirmed by qRT-PCR. Given that symptoms in COVID-19 patients take time to develop and worsen, we expect that symptom duration was shortest in patients in long-term care home and correctional facilities, longer in patients admitted to hospital ward rooms, and longest in patients residing in intensive care. Actual or approximate air change rates were available in 85 rooms. The mean air change rate/hour was 12.6 in ICU rooms, 7.5 on ward rooms, and 3.8 in long term care rooms. Estimated air change rate varied from 2 to 16 air changes/hour (mean 8.4, standard deviation [SD] 4.4).

In all, samples were positive for viral RNA in 15 (10.9%) of rooms sampled, though no viable virus was detected in any air samples. Among samples we considered positive for our analyses, Ct values for N protein ranged between 30.17 and 37.96, with a mean of 35.53 (n = 11, SD 2.06) (supplemental Table 1). For the E protein, Ct ranged from 27.03 to 36.89, with a mean of 33.61 (n = 15, SD 2.33) (Table 2). Mean RNA copy numbers for the E protein was 941.6 copy numbers/mL (range 61.3 – 11,462; standard error [SE] 752.4) and mean RNA concentration in the air was 1202.4 copy numbers/m^3^ (range 63.8 – 11939.9; SE 977.2). The highest concentration was observed in a long-term care room, though positive samples generally showed low air concentrations.

**Table 2a.**
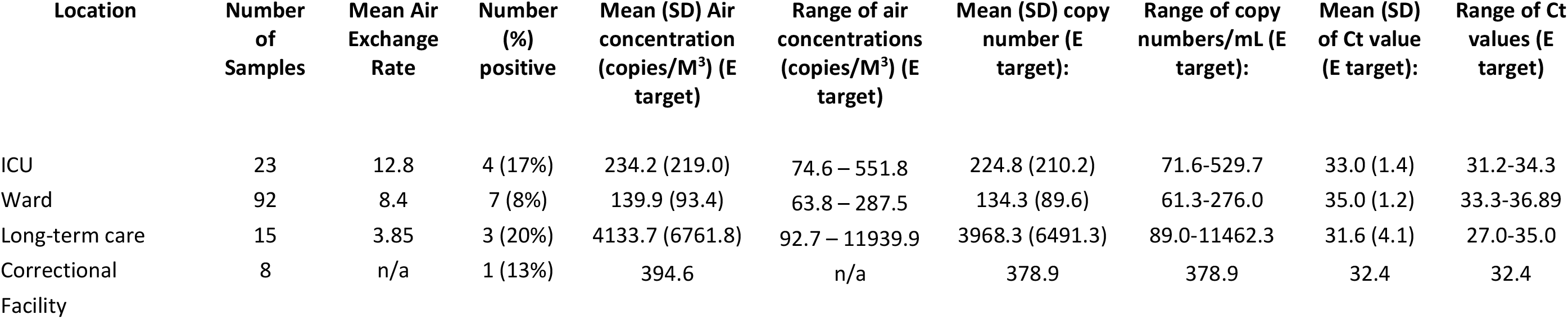
Positive SARS-CoV-2 RNA samples by room type, including Ct values for E target.

**Table 2b.** Positive SARS-CoV-2 RNA samples by room type, including Ct values for N target.

| Location | Number of Samples | N target: mean Ct (SD) for positive samples | N target: Ct range for positive samples |
| --- | --- | --- | --- |
| ICU | 23 | 35.7 (1.4) | 34.3-37.1 |
| Ward | 92 | 36.4 (0.4) | 36.0-37.0 |
| Long-term care | 15 | 34.3 (3.9) | 30.2-38.0 |
| Correctional Facility | 8 | 35.5 | 35.5 |

RNA samples were positive in 17.4% of ICU rooms (4/23), 7.6% of ward rooms (7/92), 20% of long-term care rooms (3/15) and 12.5% of correctional facility rooms (1/8). There was no significant difference in positivity rates between the different locations (Pearson Chi-Square 3.33; p = 0.34). The mean concentration of the E protein in the air was 234.2 (SD 219.0; range 74.6 – 551.8) copies/m^3^ in ICU rooms, 139.9 (SD 93.4; range 63.8 – 287.5) copies/m^3^ in ward rooms, 4133.7 (SD 6761.8; range 92.7 – 11939.9) copies/m^3^ in long-term care rooms, and 394.6 copies/m^3^ in one correctional facility return duct. We also sampled the cold air return that serviced all the inmate’s cells and living areas twice, and did not find viral RNA.

RNA samples were positive in 9.1% (6/66) of samples obtained with the UPAS 2.5 samplers, 13.5% (7/52) with the UPAS 10 samplers, and 10.0% (2/20) samples obtained with the Coriolis samplers (Table 3). There was no significant difference in positivity rates with the different samplers (Pearson Chi-Square 0.59, p = 0.74). However, sampling results should not be directly compared between the UPAS and Coriolis, because they were deployed for very different durations. A logistic regression was performed to ascertain the effects of room and sampler type and estimated air change rate on the likelihood that a positive RNA result would be observed. The logistic regression model was not statistically significant (Table 4; p value for each independent variable > 0.15). There was no significant different in E protein copy number, Ct value, or copy aerosol concentration in the various room types (data not presented).

**Table 3.** SARS-CoV-2 RNA sample results by location, sampler type

| Location | Total number of Collection events | Sampler | Number of samples by sampler type (n) | Number (%) Positive by sampler | Mean Ct for positive samples (E target) | Range of Ct for positive samples (E target): | Mean Ct for positive samples (N target): |
| --- | --- | --- | --- | --- | --- | --- | --- |
| ICU | 12 | 2.5 Micron | 11 | 2 (18%) | 32.75 | 32.75 | 37.06 |
|  |  | 10 Micron | 1 | 0 | n/a | n/a | n/a |
|  |  | Coriolis | 11 | 2 (18%) | 32.5 | 31.23-33.77 | 34.96 |
| Ward | 51 | 2.5 Micron | 49 | 4 (8%) | 35.29 | 33.71-36.89 | 36.48 |
|  |  | 10 Micron | 34 | 3 (9%) | 34.60 | 33.30-35.61 | 36.23 |
|  |  | Coriolis | 9 | 0 | n/a | n/a | n/a |
| Long-term Care | 15 | 2.5 Micron | 2 | 0 | n/a | n/a | n/a |
|  |  | 10 Micron | 13 | 3 (23%) | 31.59 | 27.03-35 | 34.30 |
|  |  | Coriolis | 0 | n/a | n/a | n/a | n/a |
| Correctional Facility | 8 | 2.5 Micron | 4 | 0 | n/a | n/a | n/a |
|  |  | 10 Micron | 4 | 1 (25%) | 32.44 | 32.44 | 35.47 |
|  |  | Coriolis | 0 | n/a | n/a | n/a | n/a |

**Table 4.** Logistic Regression results showing likelihood of positive RNA by air exchange rate, room type, and sample type.

|  | B | S.E. | Wald | df | Sig. | Exp(B) |
| --- | --- | --- | --- | --- | --- | --- |
| Air Change Rate | -.039 | .128 | .090 | 1 | .764 | .962 |
| Room Type |  |  | 3.749 | 2 | .153 |  |
| Sampler Type |  |  | .238 | 2 | .888 |  |
| Constant | -1.096 | 1.141 | .922 | 1 | .337 | .334 |

## Discussion

COVID-19 infection due to the novel SARS-CoV-2 virus has caused a global pandemic, with over 165 million cases reported worldwide, and over 3.4 million deaths at the time of this writing.[43] Furthermore, respiratory morbidity, activity limitation, and mental health conditions are prevalent, among other complications.[44] Previous studies have suggested aerosol transmission is occurring beyond close contact, as per evidence from super spreading events, transmission occurrences between adjacent rooms, viable virus measurements in air, and animal studies.[8] Our data suggests that SARS-CoV-2 RNA virus may be present at low levels in aerosols <10um in diameter, >2 m from COVID-19 patients in a variety of settings, but viable virus appears to be uncommon, as has been described elsewhere.[6]

In classical terms, respiratory viruses have been considered to be spread by droplets. Large droplets (e.g., > 5 microns in diameter) were believed to contaminate the immediate environment of an infectious patient, including the air within 2m, leading to infection by direct deposition of virus onto mucosal surfaces. In addition, large droplets settle in the proximal (within 2m) environment, leading to fomite transmission where contaminated surfaces are contacted by another person prior to touching their face thereby acquiring infection. In classical terms, aerosol (or “airborne”) spread occurs via small droplets (e.g., < 5µm) that can remain suspended in air more than 2m from a patient, leading to infection of susceptible people, via inhalation at a distance.[45] Historically, most respiratory pathogens are thought to be spread through larger droplets and fomites, with the exceptions of tuberculosis and the measles and varicella viruses, which are known to exhibit distal aerosol spread.[46] There is also evidence that influenza virus may be spread through the aerosol route.[19,46] However, the distinction between droplet and aerosol spread is fairly artificial, whereas in actuality, aerosols and droplets may both cause transmission within close contact where they are in highest concentrations, and particles many times larger than the presumed > 5µm cut-off, and perhaps up to 100µm, remain suspended in air for longer durations and distances.[25] Moreover, inhalation of aerosols may be an important, even dominant transmission mode both within and beyond close contact.[8,23]

We sampled indoor air for viral RNA and viable SARS-CoV-2 virus in a large number of rooms in a variety of settings and with a variety of room air change rates. We were careful to always sample two or more meters from COVID-19 patients, to ensure that we were detecting virus only at distances traditionally considered to be consistent with airborne transmission. Despite this variety of indoor environments, viral RNA was detected infrequently. Only 10.9% of rooms contained viral RNA, and while detection rates were highest in ICU rooms and rooms in long-term care facilities, the differences between room types were not statistically significant. Differences in detection rates likely reflect a multiplicity of factors, including each patient’s infectivity and duration of illness, room size, and ventilation rates.[24] No viable virus was found. This likely reflects the same factors. In addition, Ct values were low, and there is good evidence that detection of viable virus is unlikely when the Ct value is greater than 24.[27] Our mean Ct values were just over and under 34 for the N and E proteins, respectively. The Ct value was under 34 for the N protein in only one room, and under 34 for the E protein in eight rooms. We did not observe a significant difference of RNA positivity rates using samplers capturing larger (10μm) or smaller particles (2.5μm) or non-sized particles, suggesting viral RNA may be present in a range of particle sizes. Within the potential limitations of our methodology, our results suggest that aerosol transmission risk beyond 2m was low in hospital rooms, at the time of our sampling. While hospitalization is more likely to occur later in the disease course, when infectivity is lower, caution is still warranted.

Expert groups have examined the plausibility of aerosol spread of SARS-CoV-2 virus to beyond 2m, with some groups reporting that such spread was likely or occurring.[5,8,45,46] However, air sampling data supporting this assertion is limited, and as such, a recent WHO-funded panel maintains that additional air sampling data is needed to draw conclusions, due to the lack of viable SARS-CoV-2 detected in air.[26] Reports in the literature present contradicting data points but all agree that there is detectable virus within aerosol samples. Liu and colleagues reported finding SARS-CoV-2 RNA in several aerosol samples in hospitals in China, including a patient toilet room and isolation anterooms where personal protective equipment was doffed, although resuspension of contaminated surfaces can’t be excluded.[11] In contrast, Cheng and colleagues did not detect viral RNA very close to patients in isolation rooms with an air change rate of 12 air changes per hour.[47] Razzini and coworkers obtained five air samples and detected viral RNA in converted negative pressure operating room air and a corridor.[48] Chia found viral RNA in two of three air samples from airborne isolation rooms in a hospital in Singapore, including particles 1 to 4 and > 4 microns in size.[30] An air exhaust vent and many room surfaces had RNA present.[30] Santarpia detected viral RNA in a biocontainment unit in Nebraska, USA in an air handling grate, 58.3% of corridor samples, and a sample at a doorway over 2m from a patient receiving oxygen by nasal cannula. Some evidence for viable virus was detected.[49] Numerous room surfaces also had viral RNA. However, in a small study of 6 hospital patient rooms in this unit, viral RNA was detected at the foot of each patient’s bed, and 3/18 samples had culturable virus, including 2 of the 1-4 micron samples.[50] Similarly, in a small study of 2 patients in hospital in Florida, USA where sampling was designed to prevent any damage to virions, Lednicky and coworkers observed viral RNA and viable virus 2 to 4.8 m away from both patients. The air change rate was 6 air changes per hour. [40] Lednicky also reported finding viable virus in particles 0.25 – 0.5 microns about 1 m from a patient with minimal symptoms, inside her car.[12] In a larger study of 20 patient rooms in North Carolina, USA where the air change rate was about 14 air changes per hour, viral RNA was found in three rooms (15%): particles < 4 microns about 1.4 and 2.2 meters from the head of the bed, and over 4 microns 2.2 meters from the head of the bed, respectively. No viable virus was detected. Hallway and clinician work stations did not have detectable virus.[51] Similarly, a study of 22 patient rooms in Quebec, Canada found viral RNA in six rooms (27.2%), but no viable virus. Rooms had a mean air change rate of 4.85 air changes per hour, and samplers were located at a window, an unspecified distance from the patient’s head.[52] These studies were generally limited by small sample sizes, inadequate description of the distance between the patient and the sampler, inconsistent methodologies and reporting, and lack of determination whether viable virus was present.[26]

The UPAS sampler was appropriate for use in this study, and provided flexibility for collecting aerosol samples in multiple environments. It is compact, lightweight and quiet, so it is easily deployable and suitable for personal monitoring. In addition, its internal pump, internal battery, and lack of protruding pieces or external tubing made it more acceptable for sampling in clinical environments, where clinicians have limited time, and would be averse to deploying samplers that may increase contamination risks or disturb patients. Its main benefit for this study was that it can be fitted with validated size-selective inlets, which allowed us to collect only respiratory particles in the size fractions conventionally defined as aerosols.

It is worth noting that aerosol sampling, and subsequent culturing, may not be a necessary indicator of whether or not a virus is spread through aerosols. Infectious and non-infectious virus are expelled by the respiratory tract via similar mechanisms, so the presence of RNA in air may indicate that aerosol transmission is possible. Indeed, recognized airborne diseases such as measles and tuberculosis have never been successfully cultured from indoor air.[8]

Our study had several limitations, which may have affected our ability to detect airborne viral RNA and viable virus. In accordance with REB requirements, no direct sampling of patients was performed to determine their infectiousness, and we did not have access to patient history, including demographics or symptoms, or illness duration. Almost all hospitalized patients were admitted at least five days after symptom onset, when they are less likely to be shedding infectious virus, though they may still shed non-infectious RNA. [24] Studies have shown that patients shed viable virus for a fairly short period of time (on average for several days before the onset of symptoms, and for 8 days or less after symptom onset), and it is possible that sampling earlier in the course of these patients’ illness would have had a higher positivity rate.[53] Also, viral shedding may not be a continuous event, even during the infectious stage. It is possible that shedding is intermittent, and our 16hr sampling may have missed these events. Furthermore, the amount of shedding varies substantially between people, and we may not have sampled near high-emitters.[6,54] Our data is consistent with observations that the window of time where infectious aerosol may be present is generally short.[53] While no viable virus was detected, we believe that if sampling was conducted earlier in the course of infection, it would be more likely that viable virus would be detected, as viable virus is present in the same size fraction as the non-infectious virus we detected, occasionally in high concentrations.[12] However, sampling was also conducted near penitentiary prisoners many of whom were asymptomatic when diagnosed or were within a day or two of symptoms; and in LTC and ward patient rooms identified through routine testing as part of an outbreak. Viable virus was not detected in those samples. Unexpectedly, higher rates of viral RNA detection did not occur during sampling in rooms in several long-term care facilities and in penitentiary cells during institutional outbreaks, despite these patients being in early stages of infection, when they were asymptomatic or had likely only recently developed symptoms. Moreover, these sampling events took place in locations where room air changes rates were either known or presumed to be lower than hospital settings. Room ventilation data was obtained by hospital facilities management departments; in many cases, these were measured directly in study rooms, but in other cases, only data from “typical” or similar rooms in the facility were available.

Several issues could have affected our ability to capture viable virus beyond close contact. The vast majority of our collections took place in well-ventilated environments, where it is less likely to find aerosols. It remains possible that the act of sample collection (i.e., initial path of virus through size-selective inlet and into sampler) could reduce virus viability, as we could not test this experimentally. However, Lednicky et. al., successfully captured viable SARS-CoV-2 using a similar size selective inlet in similar conditions.[12] Also, if the samplers led to fragmentation of virus, we should have observed relatively large quantities of fragmented RNA, which was not the case. To mitigate this risk, we chose a gelatin filter to collect any virus drawn into the samplers, to maximize viability of the virus. We confirmed through method validation experiments that SARS-CoV-2 viability would be preserved on the gelatin media. Given the low Ct values of viral RNA we observed, it was unlikely we would observe viable virus, as has been observed elsewhere.[13,55] Specifically, Bullard et al. reported that no viral growth was observed when the Ct was greater than 24.[24] It is possible that our airborne sampling degraded virus making the detection of viable virus unlikely, but we confirmed that our samplers preserved the viability of a surrogate virus, and if we captured viable virus that had disintegrated during capture in our samplers, we would expect low Ct values reflecting large amount of fragmented viral RNA.

## Conclusions

This study carefully assessed whether infectious SARS-CoV-2 virus is present in indoor air at distances felt to be compatible with aerosol transmission of virus. In generally well-ventilated spaces, we found only low concentrations of viral RNA at >2m distance, and we did not detect viable virus. Concentrations in the air averaged 1202 copies/m^3^ and ranged up to 11939 copies/m^3^. Previous studies found viable SARS-CoV-2 aerosols in samples with similar RNA concentrations.[12,40]

Restricting the sampling to the <2.5µm and <10µm size fractions ensured that only viruses present in aerosols were captured, and provided additional information on the size fractions containing the virus. Together with the field data collection, our method development experiments demonstrate that conventional air sampling equipment (i.e., personal air sampling equipment) may be used for the purpose of virus sampling, with the support and expertise on particle capture from the aerosol sciences, and lab analysis and interpretation from virology and medicine.

The information present here on levels of virus in respiratory particles, their presence in aerosols >2 metres from patients, and their size, may support infection control measures and PPE guidelines in buildings preparing for re-occupancy, medical facilities, long term care facilities, and other common spaces. Our data are consistent with existing public health guidance to maintain physical distance, wash hands, wear high quality and well-fitted masks, avoid crowded and confined indoor spaces, and ensure indoor spaces are well-ventilated.

## Data Availability

All relevant data are within the manuscript and its Supporting Information files.

## Acknowledgements

We would like to thank Dr. Kathryn Suh from The Ottawa Hospital, and Lynn Kyte from the Children’s Hospital of Eastern Ontario Research Institute for their support, and Health Canada and the Public Health Agency of Canada for funding support.

